# Hygiene Knowledge and Practices in the Lagos Wild Meat Value Chain: Cultural Influences, Regulatory Gaps, and Infrastructure Needs

**DOI:** 10.1101/2025.02.08.25321724

**Authors:** Samuel N. Akpan, Elizabeth A.J. Cook, Frank van Langevelde, Pim van Hooft, Dawn M. Zimmerman, Ralph Buij, James M. Hassell, Sherrill P. Masudi, Christian T. Happi, Anise N. Happi, Lian F. Thomas

## Abstract

Wild meat, commonly known as bushmeat is a cultural, economic, and nutritional staple food in many regions of the world, including sub-Saharan Africa. Wild meat value chains face major hygiene and sanitary regulation challenges, but only a few studies have investigated these challenges, focusing instead on market dynamics and biodiversity issues. This study examines the hygiene practices, attitudes, perceptions, and knowledge of public health risks among actors in the Lagos (Nigeria) wild meat value chain, and its consequences for food safety. We employed a mixed qualitative and quantitative study design, using in-depth interviews of key informants (n=34) purposively selected from the wild meat value chain’s hunter, wholesaler, processor, and retailer nodes. An inductive thematic approach and descriptive statistics were used for data analysis. Results revealed three overarching themes: wild meat process hygiene, personal hygiene, and governance. Social norms, distrust, infrastructural deficits, and poor regulation were the drivers of the hygiene practices in the value chain. Actors showed poor knowledge of the health risks associated with wild meat, prioritizing taste over its safety. Women were more at risk of contracting zoonotic infections due to gender biases which exposed them to riskier nodes of the value chain. Wild meat as a food system in developing urban cities presents a potential source of food-borne disease transmission and zoonotic pathogen spillover to actors and the general public, due to underlying poor hygiene practices. We recommend intervention approaches that integrate peoples’ cultures, provision of infrastructure, enforcement of sanitary standards, actors’ education, and further empirical research to stimulate the establishment of hygiene guidelines for the regulation of urban wild meat value chains.

## Introduction

Wild meat—commonly referred to as bushmeat—holds significant global cultural importance, including in sub-Saharan Africa [1]. For example, the commercial city of Lagos in Nigeria serves as a major hub, supporting a thriving market, for both the consumption and trafficking of wild meat [2]. However, poor hygiene practices within the wild meat value chain have long been recognized [3], with low levels of awareness around meat hygiene being particularly evident among wild meat vendors in large urban areas [4]. Inappropriate handling, processing, and improper storage often lead to meat contamination, which negatively impacts both meat quality and safety, and contributes to the transmission of zoonotic and foodborne pathogens [5,6]. These challenges are exacerbated by inadequate infrastructure, weak regulatory enforcement, socioeconomic constraints, and entrenched cultural beliefs [7]. Wild meat food safety regulations are often neglected in large urban areas, either due to limited resources or corruption [8,], which directly influence sanitation practices. Moreover, cultural variations across communities play a significant role in shaping wild meat practices, including trade policy and food culture [9]. Rapid urbanization and a growing population have increased the demand for wild meat, putting additional pressure on already fragile ecosystems [10]. Historically, concerns about the wild meat trade have primarily focused on issues related to biodiversity conservation [11], but the high-profile disease outbreaks linked to wild meat and contamination of food by wildlife in urban areas, has shifted the focus to public health concerns. In 2017, a deadly outbreak of zoonotic Lassa fever emerged in Lagos and nearby countries in West Africa [12]. The virus was linked to rodents, particularly the multimammate rat (*Mastomys natalensis*), which played a crucial role by indirectly transmitting the infection to humans through contaminated food products [13]. The 2017 outbreak of Ebola virus disease (EVD) in the Likati zone of the Democratic Republic of Congo was linked to a hunter who had contact with fresh wild meat (monkey and wild boar), while the first Lassa fever cases in Ghana’s Ashanti region were traced to persons who had consumed rodents [14,15]. Despite these health risks, the wild meat trade continues to flourish due to cultural and economic factors [9]. Proper wild meat hygiene is essential to minimize the risk of contamination and transmission of zoonotic diseases. According to [16], unhygienic handling of meats by commercial vendors and processors accounts for 79% of food-borne disease outbreaks. To support food-borne and zoonotic disease risk mitigation strategies, it is imperative to deepen understanding of the hygiene and handling practices within the wild meat value chain, as well as determine the knowledge and attitudes of those involved in the trade, particularly concerning food safety and public health.

In this paper, we focus on Africa’s most populous megacity—Lagos, Nigeria—where only limited research on this subject has been conducted. The lack of empirical investigation into wild meat hygiene in this region means that the health risks posed by the Lagos wild meat value chain may be underappreciated. This study identifies the major hygiene and sanitary regulation challenges of urban wild meat value chains, providing crucial data to support interventions that aim to improve global food safety, enhance environmental health, and strengthen hygiene policy enforcement in Lagos and other urban cities with similar challenges in sub-Saharan Africa and other regions of the world.

## Materials and Methods

### Study area

This study was conducted in Lagos, situated in southwestern Nigeria. Lagos’s dense population, estimated at 26 million people [17], supports a bustling center of commerce where wild meats are traded alongside other goods. Figure 1 presents a map of Lagos city showing the local government areas and wild meat value chain nodes.

**Figure 1.**
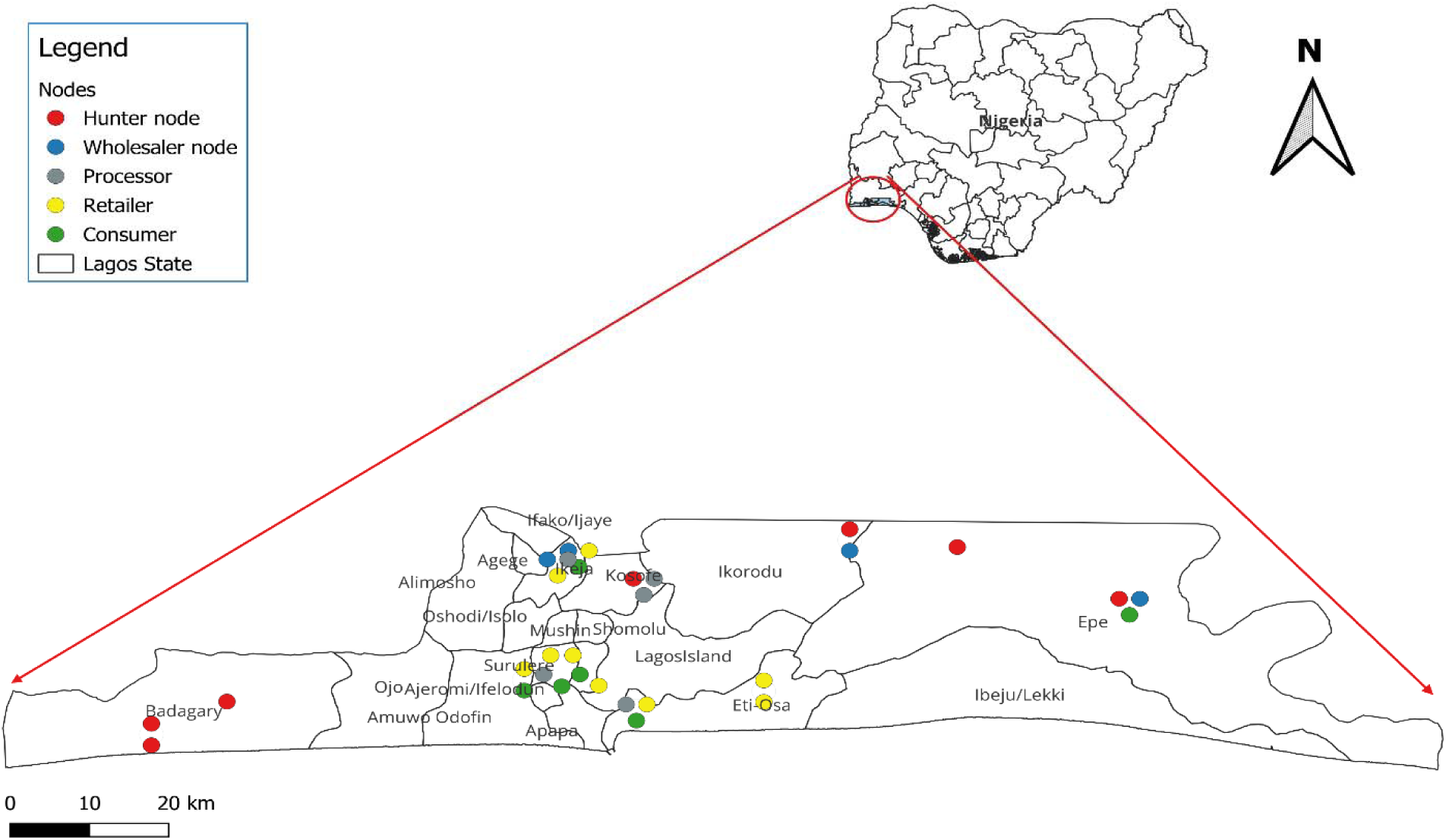
Local government areas and wild meat value chain nodes in Lagos, Nigeria.

### Ethical approval

Approval for this research was received from the National Health Research Ethics Committee (NHREC) of Nigeria, with approval number NHREC/01/01/2007-07 /12/2022.

### Ethics statement

Human study participants were recruited from 22-01-2023 to 17-06-2023. Before every interview, the research team shared the project information with the participants through a Project Information Sheet (S1) and sought their consent to participate in the study (S2). We received verbal consent from all participants in this study. Each consent was documented by ticking on the consent form with the initials of each participant. All consents received were physically witnessed by the lead author.

### Data collection

The data presented in this manuscript relates to items 8–18 of the interview guide (**S1**). Semi-structured interviews were conducted and geographic coordinates were captured during a Lagos wild meat value chain mapping exercise. Participants were purposively selected based on their roles in the value chain. They were hunters (n = 8), wholesalers (n = 5), processors (n = 9), and retailers (n = 12). Although the consumer nodes were identified, no wild meat consumers agreed to participate in the study. With the aid of a local interpreter, interviews were conducted in the local Yoruba language for participants who could not understand or speak English. All responses were recorded. We asked the participants about their roles in the value chain, wild meat regulations, and their hygiene practices. “Hygiene” in this study context was used to refer to the specific methods and practices used by individuals involved in the hunting, processing, transporting, storing, preservation, packaging, displaying, and selling of wild meat which may impact its quality and safety. Participants were also asked about their perception of zoonoses, and if they believed wild meat may be a source of zoonotic and food-borne disease transmission to humans. To gain more insight on how to improve the hygiene situation, we sought to unravel the motivations behind the reported hygiene practices, using the transcribed data. This was mostly indicated in follow-up questions which had to do with “why” actors might do the described activities.

### Data analysis

Descriptive statistical analysis was conducted on quantitative data using Microsoft Excel. Recordings from the interviews were transcribed verbatim into Microsoft Word and analyzed qualitatively by thematic analysis, using an inductive approach. The analysis process involved familiarization, coding, theme generation, reviewing, defining, and reporting [18]. The transcripts were read multiple times by the researchers to review any discrepancies between the audio recording and the written records of the responses from the participants. From the familiarization, the initial codes were arrived at using the study themes. From the initial codes, major codes emerged in the forms of words, phrases, and sentences, as described by [19]. Systematically applying these codes to the entire dataset led to the discovery of patterns, connections, and recurring themes. This process aided in theme formation through the grouping of similar codes. Categories already covered were collapsed. Through this process, the major codes evolved into themes. To ensure that the major themes captured the essence of the data, the initial themes were reviewed and refined by comparing them with the coded data. The themes that captured the essence of the data they represent were retained while those that deviated were either refined, restructured, or merged. We considered the factors (emergent codes) grouped under each of the three overarching themes as drivers of the hygiene practices of actors in the value chain. **Table 1** shows the overarching themes, codes, and their description.

**Table 1.**
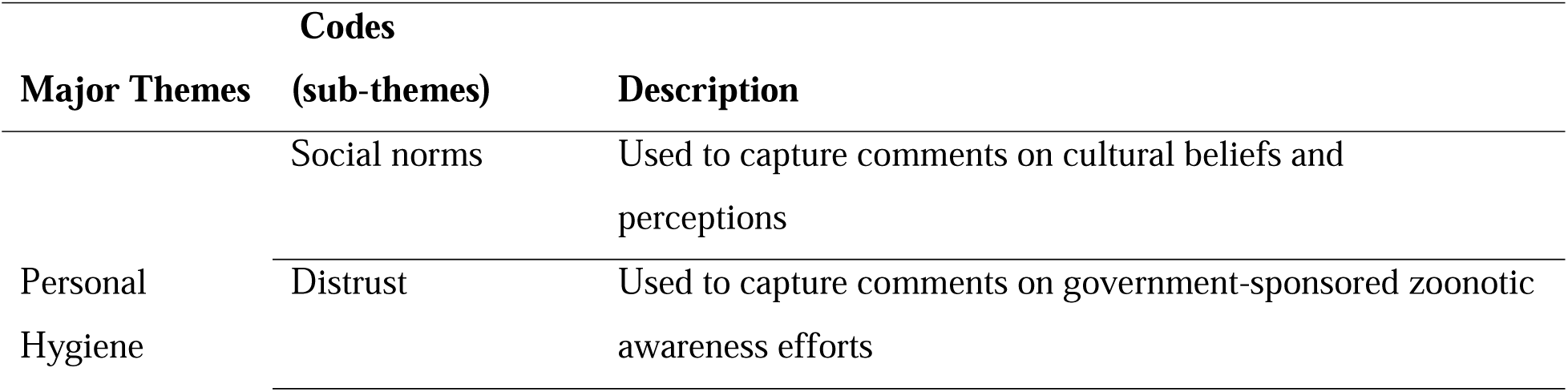
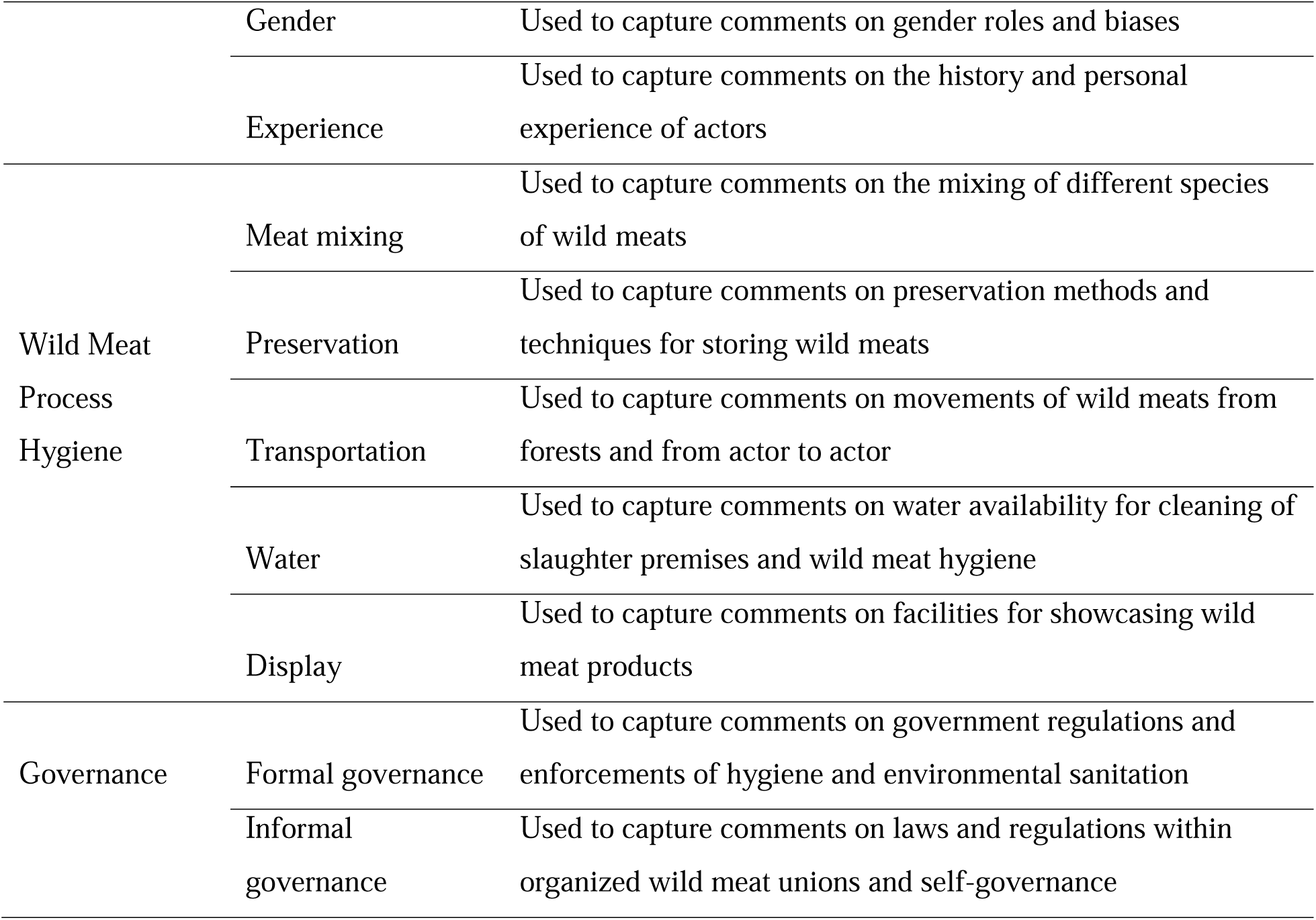
Codebook showing the emergent codes and their covering themes Codes.

## Results and discussions

This study identified key issues for the wild meat value chain in Lagos, Nigeria, each presenting unique challenges as a result of social norms, practical constraints, and infrastructural challenges. We believe these are representative of other sub-Saharan countries, as countries in this region are known to have established robust wild meat markets and traditions [20,21,22].

### Participants’ roles in the value chain

Diverse but complementary value chain roles and activities were reported. This encompassed sourcing/production, processing, packaging, sales, and marketing, as shown in **Table 2**.

**Table 2.**
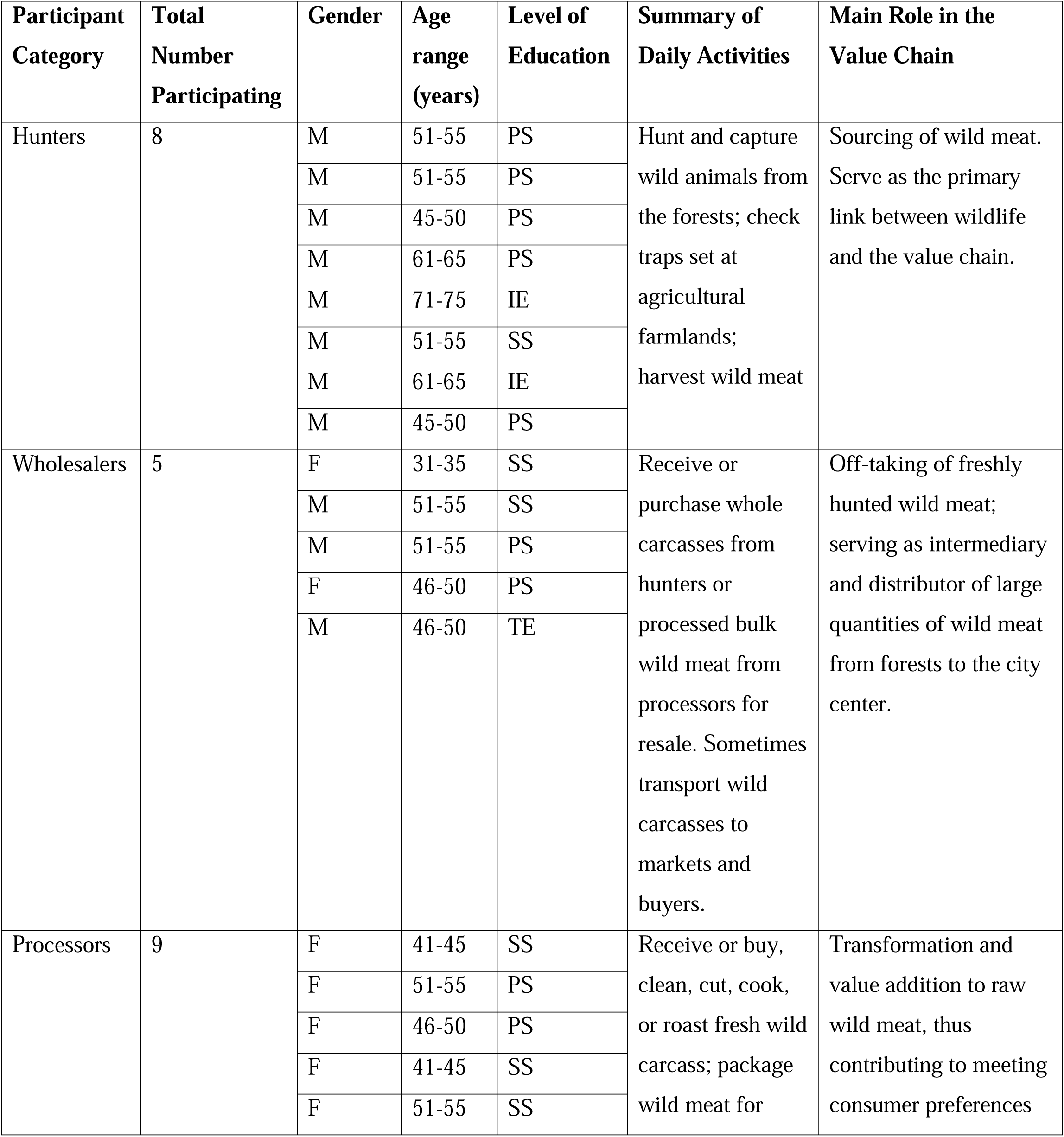
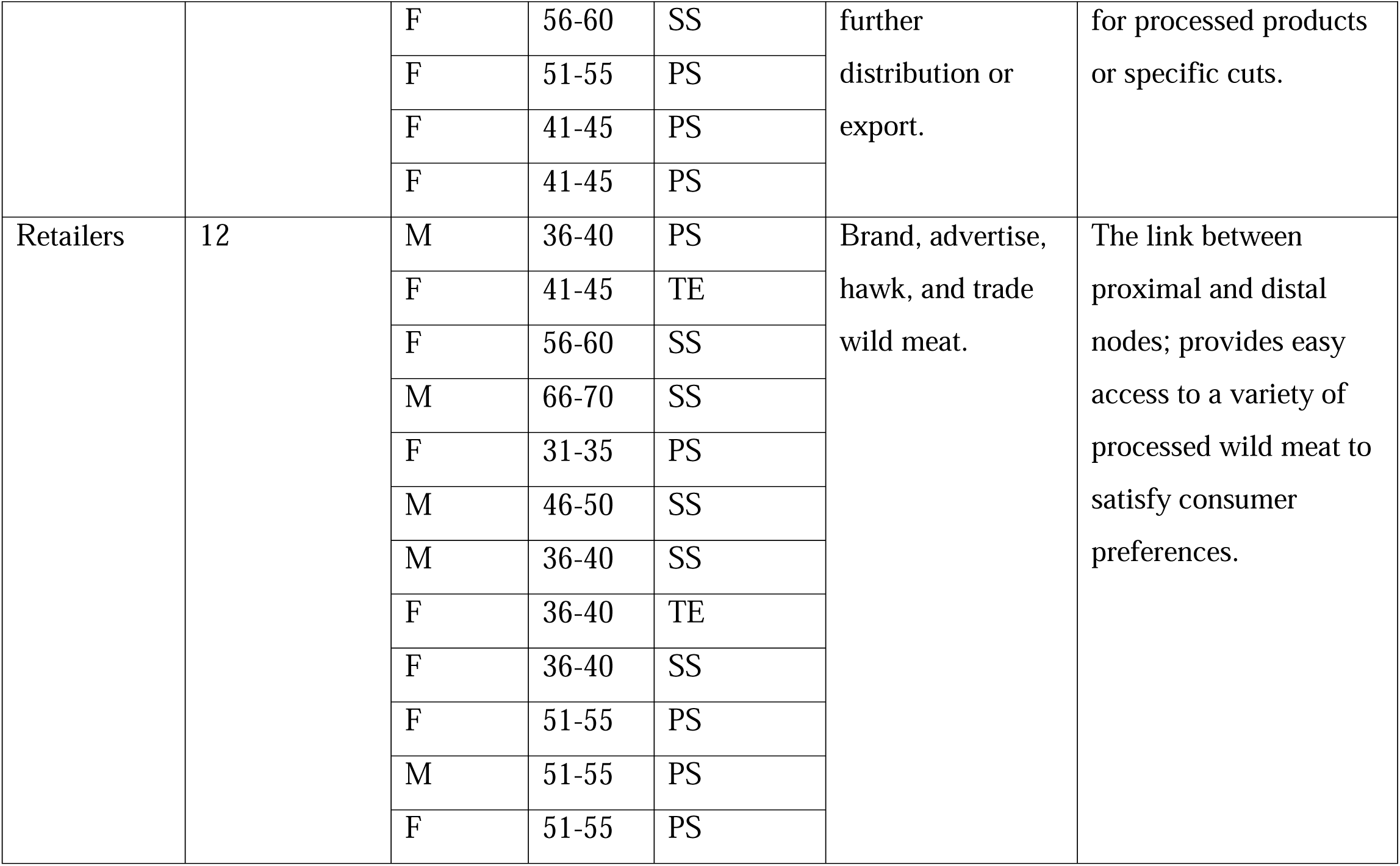
Participants’ Demography and Roles in the Value Chain.

Half of the participants (50%) had attained primary school education, 35.2% had secondary education, and 8.8% and 5.8% attained tertiary education and informal education, respectively. The majority were females (53%), and males accounted for 47%.

### Hygiene practices of actors in the Lagos wild meat value chain

Results of the thematic analysis revealed three overarching themes: (i) personal hygiene (ii) wild meat process hygiene and (iii) governance. A graphical representation of the participants’ responses is shown in Figure 2.

**Figure 2.**
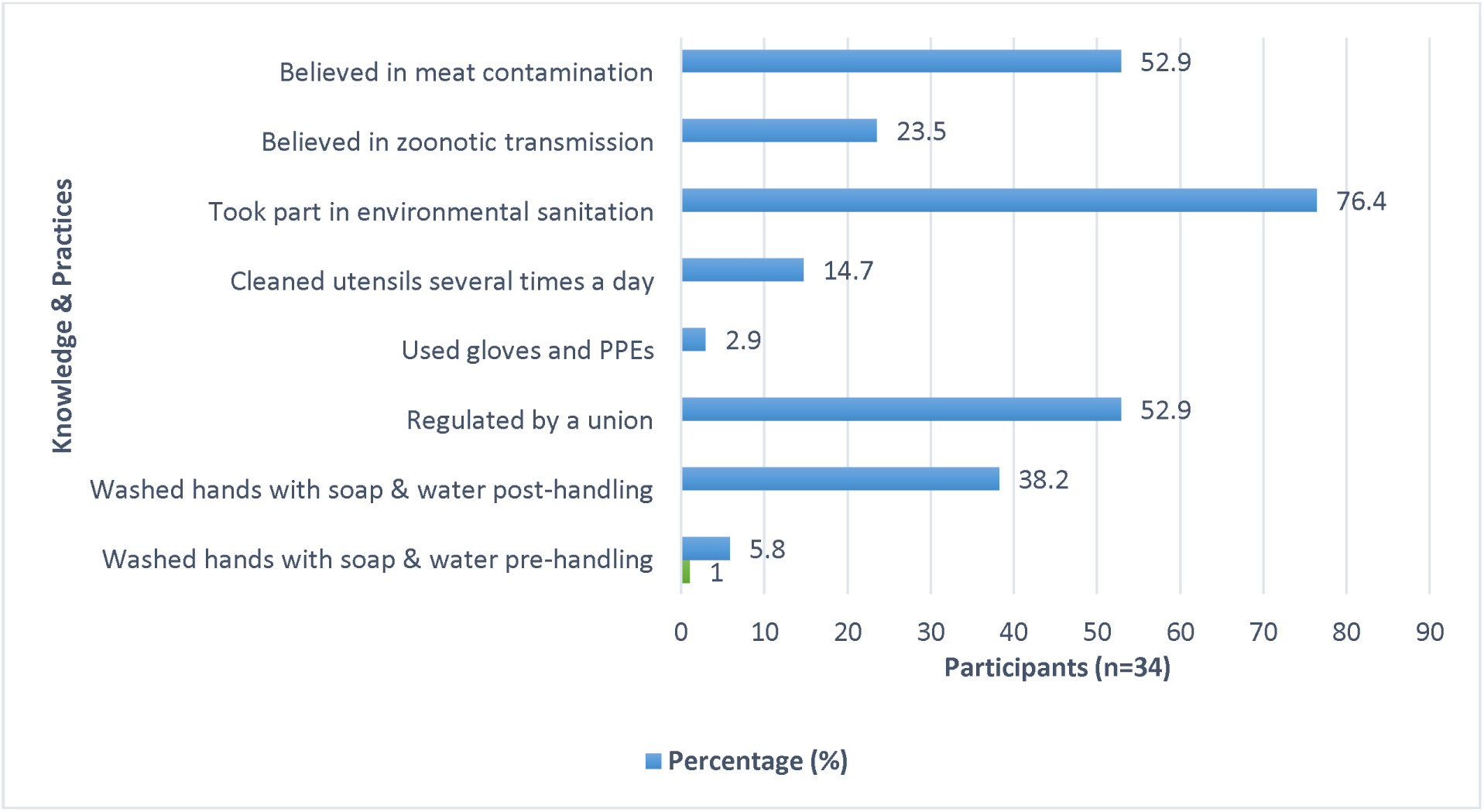
Frequency chart of participants’ responses on their hygiene knowledge and practices in the wild meat value chain.

### Personal hygiene

Here we describe the practices that were reported by participants or observed by the researchers during data collection, which we see as being explicitly linked to the wild meat actors’ hygiene. Four sub-themes emerged under this major theme. They were: social norms, distrust, gender, and experience.

### Social norms

Cultural beliefs and perceptions dictated certain wild meat handling practices among actors. For example, actors did not prioritize hand washing with soap after handling fresh or processed wild meat, because they believed they were immune to infectious diseases:

> *“Washing hands with soap after handling is not so necessary because as black people, our immunity is very high, hence we can’t be sick [through wildmeat]. Take the case of COVID-19 for example”* (Wholesaler).

Another common belief was that wild animals were pure and hence could not transmit diseases:

> *“………I don’t need to wash my hands after handling, because wildlife is pure. There is nothing like that kind of disease [zoonoses]”* (Processor).
>
> *“After handling, we rinse hands with whatever we are drinking, especially beer or palm wine. I hear it is even better than hand sanitizer”* (Hunter).

Participants’ observation revealed some actors ate with bare and unwashed hands while also processing wild meat.

Hand hygiene is a critical factor in the safety of final meat products. Hence, hand washing and the use of gloves have been outlined as the main methods for reducing bacterial cross-contamination from hands to ready-to-eat food [23]. According to a study conducted by [24], food workers’ hands harbored food-borne pathogens (*Escherichia coli* and *Staphylococcus aureus)*, which could potentially contaminate meats sold for public consumption. Also, [25] report that washing hands with soap before and after handling food products can lead to a marked reduction in the probability of reported food-borne illnesses. While there is no documented evidence of the harm of hand washing with palm wine, beer, or other alcoholic drinks, the practice is inappropriate and a breach of established food safety standards. Actors also face the risk of contracting zoonotic and food-borne infections arising from cross-contamination, when they eat and feed their young ones with unwashed hands from wild meat processing. Unfortunately, consumers are the main victims of health concerns related to handling and equipment as these unhygienic practices lead to food poisoning resulting from microbial contamination [26].

Traditional hunting practices were also exhibited in the handling of wild meat. Fresh carcasses were carried on shoulders, while some were butchered in the forests:

> *“Anytime we get [hunt] rodents, we put inside a sack bag; but for animals like antelope or deer, we carry it on our shoulders. I don’t know why but that is how we do it.”* (Hunter).
>
> *“When I get wild animals, I butcher and roast it there on the farm before taking it home”* (Hunter).

The excerpts indicate that certain risky meat hygiene practices were borne out of age-long traditional practices and regarded as norms by actors. This is corroborated by [27], who stated that the predisposition to process meat in unsanitary conditions is often driven by cultural practices and the lack of simple technology as an alternative (eg carts and wheelbarrows). Social norms not only hinder the adoption of proper hygiene practices but sustains risky behaviors among actors in the value chain. Similar to our findings, [7] reported a significant influence of cultural factors on food safety practices due to traditional beliefs and practices that preceded scientific recommendations. To buttress our finding, [28] observed that culturally influenced practices were majorly practiced and perpetuated in the Lagos wild meat value chain through hunters historically regarded as custodians of the Yoruba culture of the people of Southwestern Nigeria.

### Gender

Gender roles influenced hygiene, as the men often avoided tasks perceived as women’s work e.g., cleaning and processing. Hence, women were responsible for processing and preparing wild meat delicacies at processing and retail points, drinking bars, and homes:

> *“As a farmer, I set traps in my farm, and sometimes I am lucky to catch some rats and other animals. When I return home, I usually hand them over to my wife to butcher and prepare for the whole family to enjoy”* (Hunter).
>
> *“……I don’t process bushmeat, because it is the duty of our women. The only challenge is that sometimes they may be few and before they can finish, some animals [carcasses] may start to rot because of the long period.”* (Hunter).

Nursing mothers were also observed to juggle the task of caring for their young with wild meat processing.

The outlined gender roles limit men’s involvement in tasks considered to be within the domestic domain, such as cleaning and processing. Such scenarios can lead to gaps in hygiene practices, especially in contexts where there are not enough women to perform these tasks. Hence, the reluctance by men to engage in cleaning and processing activities perceived as women’s tasks is a critical barrier to achieving overall environmental hygiene conditions in wild meat markets, processing, and retail points. As reported by [29], the highest risks of disease transmission in the wild meat value chain occur during processing (cutting, skinning, etc). This infers that the higher involvement of women in wild meat processing creates a gendered difference in zoonotic exposure risks since women are put at a higher risk of contact with pathogens from potentially infected fresh carcasses.

### Distrust

A significant gap in the knowledge of zoonotic disease transmission through wild meat was seen. Participants in their responses were dismissive of the health risks, citing a plot by the government to disrupt their livelihoods, as captured in the following excerpt:

> *“I usually hear the government people talking about it. Let me tell you, there is nothing like zoonoses and we cannot contract any disease through handling wild meat”* (Hunter).
>
> *“That Ebola or COVID-19 thing is a lie by government people, to discourage us from our business, and discourage people buying from us. Here, we don’t believe [such]. There is nothing like Ebola”* (Processor).

These responses point to public distrust of formal institutions and the government, with actors having negative perceptions about the occupational disease risks communicated to them. This finding resonates with the findings of [30] who reported that informants in Guinea put out various explanations to explain the ban on wild meat trade during the outbreak of Ebola virus disease (EVD) in 2016; such as the government trying to bolster its hold on power by undermining villages in regions where the opposition party was supported. A study by [31] also revealed that people rejected the government ban on wild meat, and health messages linking wild meats to Monkeypox in Nigeria. If left unaddressed over time, distrust can hamper the effectiveness of risk communication and disease prevention efforts, predisposing wild meat actors and community members to zoonotic and food-borne diseases. This could also play a role in maintaining the cycle and transmission of novel and emerging pathogens both nationally and internationally.

### Experience

Results indicated that the attitudes and perceptions cultivated over time through the personal experiences of actors discouraged their use of protective clothing and equipment. They believed that using gloves would suggest their wild meat products were unsafe, which they believe will send the wrong message to intending customers, as seen in an excerpt:

> *“You should not be seen wearing hand gloves while holding wild meat. If you do, then you are telling the people that your wild meat is not safe, and they will not buy it”* (Retailer).
>
> “…… We *don’t use hand gloves……I have never seen anyone doing that. The only problem is that sometimes we get a few skin scratches from some animals that are not fully dead. But it is normal for us”*. (Hunter)

The use of personal protective equipment (PPEs) such as gloves and face masks were not practiced among the actors. Physical evidence combined with participants’ responses indicate that there was a social stigma associated with the use of protective gear by actors. However, several studies report that bare-hand contact with meat can introduce foodborne pathogens, which could lead to illnesses especially since most foodborne pathogens (e.g., *Salmonella* spp., *Campylobacter* spp., *Listeria* spp., *Escherichia coli*, etc.) have low infective doses [25,32]. Associating the use of hand gloves with unsafe meat is a pointer to the low level of awareness and poor perception of proper hygiene measures by actors.

### Wild Meat Process Hygiene

In this section, we describe particular practices that were reported by participants or observed by the researchers that could be explicitly linked to the process of wild meat harvesting, transport, and processing. The sub-themes that emerged here are: meat mixing, preservation, transportation, display, and water.

### Meat mixing

Field observation revealed the mixing of different species of wild meats. As a general practice, freshly killed animals of diverse species and sizes were mixed during transportation. Participants’ responses also indicated that the local people liked the taste and aroma that some species give to other meats when mixed during cooking or storage:

> *“After purchase, I put all the animals together into sacks, and transport them on my motorbike to the market for sales*” (Wholesaler)
>
> *“I mix antelopes, porcupines and other animals [species] with snakes during cooking. The reason is that snakes give a delicious taste to the other meat, which many of my customers love”* (Retailer).

Similarly, actors referenced consumer preferences and past generations’ practices to justify the actions:

> *“I mix different meats like rats and snakes while cooking. My customers have never complained, and I do not want to lose them”* (Retailer).

Here, the influence of consumer preferences cannot be overlooked, as there was an overbearing mindset of “customer is king”, a perception that whatever is acceptable to consumers should guide the wild meat practices. According to [33], consumers play a pivotal role in food safety as their preferences and perceptions significantly influence food production, distribution, and consumption practices. While consumer pressure derived from their risk perceptions may influence a change in meat handler’s poor hygiene practices [34], its absence suggests a gap in consumers’ food safety awareness levels [35]. This suggests that the practice of meat mixing in the value chain were also shaped by consumer ignorance.

The practice of mixing different species of wild meat and body parts (e.g., intestines, hides) to enhance taste and aroma indicates that the consumers elevated the tastiness of wild meat products over their safety. This practice can lead to cross-contamination of meats with bacteria or other viruses, resulting in food-borne illnesses to the consumer. Also, the close contact of different species of freshly killed animals in wildlife markets can be an avenue for the potential re-assortment and evolution of many zoonotic viruses which previously could not affect humans [36].

### Preservation

The lack of refrigeration infrastructure and equipment for preserving and cooking wild meat during hunting expeditions also influenced the practices:

> *“Sometimes there are poor sales and I still have some meat [fresh carcass] which I don’t want to spoil [rot] and produce a bad smell. So, I usually remove the intestines and other organs, to slow down the spoiling [spoilage]”* (Wholesaler).
>
> *“While hunting, I sometimes eat stale or half-cooked meat. Because the forest is [may be] wet so the wood cannot burn to heat up or roast any meat very well”* (Hunter).

The lack of basic infrastructure such as preservation facilities often limits actors’ adoption of good hygiene practices. Poor meat preservation can lead to the growth and proliferation of bacteria which cause meat spoilage. In many tropical developing countries, meat spoilage is often caused by a lack of storage facilities and unfavorable ambient temperatures [37]. Also, certain environments present limitations for meat hygiene, especially in low-resource settings. For example, due to poverty levels, it may be difficult for hunters to afford access to refrigeration facilities to keep and or preserve their meat after hunting expeditions. Worthy of note is the lack of energy needed to power refrigerators and cold chain facilities. [38] report that over 760 million people worldwide lack access to electricity, especially in rural communities of South Asia and sub-Saharan Africa. This may be the underlying factor driving the poor meat storage and preservation practices seen in this study.

### Transportation

During hunting expeditions, large animals such as buffaloes are slaughtered in the forests, where sanitary hygiene is not guaranteed:

> *“When we shoot a large animal, we butcher it in the forest so that transporting it will be easy”* (Hunter).
>
> Due to transportation constraints, meats from different species were mixed. Different body parts (e.g., hoofs, horns, intestines, and hides) were also mixed with meat cuts, increasing the risk of cross-contamination:
>
> *“As you can see, I put all the animals inside my hunting bag; both grasscutter, hare, antelope, rats, and others. That way, it is easy for me to carry them out of the bush”* (Hunter).
>
> “*During group hunting, we slaughter the animals in the forest, share the parts amongst ourselves, and put all of them together inside our hunting bags*” (Hunter).

As reported by previous studies, wild meat hunting in Africa is predominated by persons of low-income levels [39,40]. Hence, they are not able to afford hunting vehicles and trucks to convey their harvests from the forests. Also, ancient social norms used for team cohesion during group hunting [28], eg. sharing of meats in the forests, ensure that poor meat handling practices are maintained, predisposing meats to contamination. These practices also expose the hunters to occupational hazards such as zoonotic infections arising from direct contact with wildlife in unhygienic environments.

### Water

Field observations revealed that processors washed fresh wild carcasses with water drawn from artesian water wells, boreholes, nearby rivers, or rainwater collected in containers, depending on the availability of these sources. Others paid for traditional water vendors popularly called “*Mai ruwa*” to supply them with water sourced from farther places. Also, processors did not wash knives frequently due to the lack of water, as illustrated in the excerpt below:

> *“I don’t need to be washing my knives all the time. In this place that you see us, where will I get the water to do all that? I wash everything when I am done for the day”* (Processor).

### Display

Field observations revealed the lack of requisite facilities for safe display of processed products also hampered the hygiene standards. Wild meats were openly displayed in trays or hung with ropes on roadsides with heavy vehicular traffic and movements.

> *“I put all my products here in the open so that customers and people traveling can easily see it. If I had a showcase with glass, I would have used it……although that is not popular here”* (Retailer).

The lack of requisite infrastructure significantly influenced wild meat hygiene practices. According to [41], storage of meat at 4 _LJ_C within 4 hours post-slaughter, or freezing at -20 _LJ_C is necessary to slow down the microbial growth of spoilage bacteria, ensuring that meat is preserved. However, [5] found that many countries in sub-Saharan Africa mostly rely on primitive methods of meat preservation in general (i.e., not just wild meat) due to the absence of modern refrigeration facilities. Our study results affirm this, as the lack of refrigeration equipment compelled actors to resort to rudimentary methods for wild meat preservation, such as the use of slow-burning firewood. Significantly, it is observed here that this traditional method, though resourceful and meeting consumer preferences, does not adequately prevent microbial growth. Although not an infection risk, roasting of meats also poses substantial food safety risks due to carcinogens such as polycyclic aromatic hydrocarbons (PAHs) evolving from this method [42,43,44]. Infrequent washing of knives and other tools due to water scarcity contributes to cross-contamination and the proliferation of pathogens. Hence, the lack of consistent access to potable water posed challenges to good personal and wild meat process hygiene in the value chain.

Traditional hunting practices of butchering wild animals in the forest after capture also highlight practical difficulties in wild meat hygiene. We posit that these practices were borne out of necessity, since they occurred in environments that lacked the necessary sanitation. There was also no report or observation of meat inspection in this study, inferring wild meats were not assessed for their quality and safety pre- or post-slaughter. The role of meat inspection is to protect public health through disease detection and reporting. During this process, diseased animals/carcasses are removed from the food chain. Hence, the absence of meat inspection increases the risks of zoonotic and food-borne diseases in the value chain and contributes to negative outcomes for public health. Moreover, the interconnectedness of nodes within the value chain ensures that zoonotic and foodborne pathogen contamination from the forests can be unnoticeably transferred down the chain to the consumption level.

### Governance

Here we describe particular practices that were reported by participants or observed by the researchers that could be explicitly linked to the regulation of the value chain and actors’ activities.

### Formal governance

Participants’ responses indicated that there was a compulsory environmental sanitation by-law enacted by the government of Lagos State, to which all markets, stores, and business premises were subjected. The act, referred to as “Lagos State Environmental Management Protection Law 2017” was the only formal regulation that guided the practices of wild meat actors in Lagos. Part VI (section 188) of the enacted law which established the Lagos State Environmental Protection Agency [45] states:

“*Housing estates, hotels, commercial facilities, waste management facilities, hospitals, abattoirs and livestock shall not discharge or cause to be discharged any trade and industrial effluent into the public drain or natural environment without a permit from the Agency. (2) Effluent discharged under this section shall not exceed the permissible limits/levels contained in the Regulation of the Agency*”.

> *“…… We have cleaning of our market every Thursday morning, and it is compulsory for all of us. On that day, we can only open our business at 10 a.m.”* (Retailer)

Within main city centers, environmental sanitation officers made rounds to inspect facilities such as markets, restaurants, and shops. Levies are also collected for environmental sanitation and waste collection, with fines placed as punishment for non-compliance:

> *“Although anybody can clean at any time, our general cleaning is on Thursday, and it is always a must [compulsory]. Anybody that does not do it will pay a fine, which sometimes is heavy. But that is only when the government officers come for inspection of the environment”* (Processor).

However, these are mostly possible in areas that are near the city centers, which may be attributed to the ease of accessibility to these areas, and insufficient personnel. Participants were not aware of any other formal regulations on wild meat hygiene and safety, outside the routine weekly environmental sanitation. In their view, participants believed that the inspections were ineffective, mainly focused on the city centers and collection of fines from defaulting traders. Hence, while the general cleaning exercise may be carried out whole-heartedly by some, some participants expressed that they cleaned their work environment out of compulsion, and to evade payment of non-compliance fines:

> *“The issue of cleaning every Thursday is not really for us. It is general law for all the shops and markets in Lagos. I only joined because I am part of the market. Sometimes we don’t clean, we just close business till the cleaning time is over”* (Wholesaler).

No participant reported veterinary inspection of their fresh or processed wild meat. Also, no meat inspectors were seen in the wildlife markets, processing, and retail points that were visited

by the research team. Robust revenue generation drives are a common feature in Lagos, with all business centers, markets, hotels, drinking bars, and food outlets paying environmental levies to the government. Considering that wild meats were not being inspected, this could justify the actors’ notion that the routine inspection of their premises by government sanitary officials was solely to extort monies from them, and not for ensuring adherence to good hygiene practices.

Being the primary vehicle for many food-borne diseases [46], it is crucial to evaluate meat to guarantee its safety and reduce health risks on the consumer. Modern inspection methods include comprehensive inspections at several stages, such as evaluating animals for biological risks and identifying possible zoonotic diseases, pre- and post-mortem. However, there is no veterinary inspection of wild meats in many African countries [47]. According to [48], there is a shortage of meat inspection officers in many African nations. It can also be attributed to under-resourcing, under-financing, and neglect, as personnel of relevant public food safety and regulation agencies may be oblivious of the risks posed by wild meats.

### Informal governance

There were unwritten codes of conduct guiding the wild meat association members’ activities. The wild meat association is an informal body or union comprising wild meat hunters, wholesalers, processors, and retailers in an area or community. This body is often formed and governed by members elected from amongst the wild meat value chain actors. They also form their informal regulations or codes of conduct. According to participants’ responses, only a few of these codes of conduct addressed hygiene issues:

> *“We have many laws that we create and run by ourselves. But they are mostly about the business side. The one that has to do with the meat is that no one should process or sell rotten meat”* (Processor).
>
> *“Any dried or roasted meat from the remaining from a previous day must be warmed or heated before sales on the next day. Also, no one should sell meat from an animal that died by car accident”* (Retailer).

Some participants (n=7) reported that they did not belong to any market union, did not attend any wild meat association meetings, nor could be held accountable by any established codes of conduct. Responses also indicated that in some areas, those selected for monitoring and enforcement of the informal regulations were also actors themselves in the value chain, who were often too busy with their personal businesses and schedules. As a result of this lack of oversight, wild meat from road kills and animals that died from unknown causes were occasionally supplied and processed for sale. Also, compliance with regulations was sustained by trust and the social norm that good conscience may guide members to abide by the codes of conduct, as seen in the excerpt:

> *“In our community here, we believe in conscience to guide us. There is no reason to monitor anybody on which kind of bushmeat one is selling or not. Our members know the right thing to do and are judged by their conscience, good or bad”* (Wholesaler).

Our findings indicate that the regulatory landscape governing the wild meat trade is marked by insufficient enforcement, widespread non-compliance, and the influence of cultural and economic factors. The enforcement of wildlife and food safety regulations in Nigeria is frequently undermined by corruption [8,21], which may explain the difficulties in regulating the wild meat value chain. Also, the non-affiliation of some actors to organized wild meat associations or unions as reported in this study suggests that they and others in similar positions operated without any form of regulation— constituting a huge challenge to wild meat hygiene monitoring and regulation.

## Conclusion

The wild meat value chain in the megacity of Lagos constitutes a high-risk platform for zoonotic and food-borne pathogen transmission, due to the underlying poor knowledge, perceptions, and hygiene practices of its actors. The lack of veterinary inspection of wild meats continues to exacerbate the use of unhygienic handling and processing techniques, thereby exposing wild meat actors and the public to health risks. Additionally, our study highlights that compliance with formal regulations was only practicable within organized markets, hence regulations guiding the practice of actors were mostly informally regulated. Such regulations, though derived from social norms and sustained by trust, were not sufficient to address poor hygiene practices and gaps in the knowledge of the actors. The socio-cultural resistance of actors to good hygiene practices exacerbates the difficulties in regulatory enforcement, with financial gains prioritized over occupational and public health. While traditional methods of handling and processing wild meats are culturally important, they do pose health risks to its consumers.

Consequently, we posit that educational campaigns should also target consumers to shift their attitudes towards safer wild meat practices. The perception of “*wildlife is pure*” and “*there is nothing like zoonoses*” reflects deep-seated misconceptions about the safety of wild meat. As was also emphasized by [35], such beliefs and perceptions can lead to misplaced self-satisfaction in hygiene practices, thus increasing the risks of wild meat contamination, the transmission of food-borne pathogens, and potential zoonotic spillover.

### Recommendations

As noted by [6], awareness and education of actors is key in mitigating these risks. Hence, for wild meat hygiene interventions to be effective, local cultural beliefs must be integrated. Hence, relevant global and local bodies such as food safety agencies, national and continental centers for disease control, policy makers and governments at all levels should implement culturally-sensitive education and orientation programs to raise awareness about the zoonotic and food-borne disease risks associated with poor hygiene practices, and the importance of proper handling and processing of wild meat. For heightened effectiveness, such programs should be participatory, involving community religious and traditional leaders, to ensure the message resonates with the social norms, cultural beliefs and practices of communities. Additionally, educating the consumers on the potential health risks associated with wild meat and the importance of purchasing from only accredited hygienic outlets, will motivate wild meat processors and retailers to practice good hygiene if they are to be allowed to continue in the wild meat business. The provision of hygiene infrastructure and preservation facilities can offer great marginal benefits to public and occupational health [49]. Hence, access to clean water for all actors in wild meat value chains is critical to facilitate proper sanitation and hygiene. Also, investments must be made in modern refrigeration and cold room facilities to improve meat storage and preservation. Strengthening the enforcement of existing regulations can be done through increased and equitable funding of relevant continental, regional, and national enforcement agencies. Air and land border veterinary quarantine services should be strengthened and equipped to ensure only wild meats that meet globally accepted sanitary standards are allowed passage. Standardized hygiene practices and guidelines for all actors in the wild meat value chain should also be implemented. As emphasized by [49], in areas and situations where access to clean water, soap, and personal protective equipment are limited and there are fewer options for income and nutrition, preventing meat contamination; preventing skin wounds when handling wild meats; and preserving meat via thorough cooking, smoking, drying, and curing-could help people strike a balance between food security and hygiene. Lastly, further interdisciplinary and cross-sectoral research on public health risks of urban wild meat value chains is needed to generate data that will inform targeted interventions globally.

## Data Availability

All data on regarding the conduct of this research are fully and publicly available on a data repository. DOI: https://figshare.com/s/fb85d2629c3521b85479

## Acknowledgments

The authors would like to acknowledge Ayotunde Sijuwola, Franklyn Oluwadare, John Fadele, and Oluwatobi Adedokun for assisting with the data collection; Yvonne Linton for her useful comments and edits, and Sunday Adeeko for facilitating community entry for the research team during the field work.

## Supporting Information

**S1 File:** Project information sheet

**S2 File:** Informed consent form

**S3 File:** Interview guide

**S4 File**: Thematic data

**S5 File**: Statistical data

**S6 Fig. 1:** Local government areas and wild meat value chain nodes in Lagos, Nigeria.

**S7 Fig. 2:** Frequency chart of participants’ responses on their hygiene knowledge and practices in the wild meat value chain

## References

[1] Nasi, R., Brown, D., Wilkie, D., Bennett, E., Tutin, C., van Tol, G. et al. Conservation and use of wildlife-based resources: the bushmeat crisis 2011; 9. CBD Technical Series. https://www.cbd.int/doc/publications/cbd-ts-33-en.pdf

[2] Aberoumand, A. (2014). Nutritional evaluation of some wild meats in Iran. Food Science & Nutrition, 2014; 2(3):279–285.

[3] Ntiamoa-Baidu, Y. Wildlife and food security in Africa. World Bank Publications, 1997.

[4] Nwankwo, C., & Okeke, I. Sanitary conditions and public health risks in wildmeat markets of Lagos, Nigeria. Journal of Environmental Health Science and Engineering, 2020; 18(2):301–312.

[5] Alemayehu T, Aderaw Z, Giza M, & Diress G. Food Safety Knowledge, Handling Practices and Associated Factors Among Food Handlers Working in Food Establishments in Debre Markos Town, Northwest Ethiopia, 2020: Institution-Based Cross-Sectional Study. Risk Management and Healthcare Policy, 2021;14:1155–63. 10.2147/RMHP.S295974

[6] Insfran-Rivarola A, Tlapa D, Limon-Romero J, Baez-Lopez Y, Miranda-Ackerman M, Arredondo-Soto K, et al. A Systematic Review and Meta-Analysis of the Effects of Food Safety and Hygiene Training on Food Handlers. Foods, 2020; 9(9). Available from: https://pubmed.ncbi.nlm.nih.gov/32854221/

[7] Asibey-Berko E, Agyemang OK, & Dwumfour-Asare B. (2015). Traditional knowledge and cultural values in the management of bushmeat in Ghana: A review. International Journal of Biodiversity and Conservation, 2015; 7(4):256–269.

[8] Omojokun J. Regulation and Enforcement of Legislation on Food Safety in Nigeria [Internet]. Mycotoxin and Food Safety in Developing Countries. InTech; 2013. Available from: 10.5772/54423

[9] Trefon T. Bushmeat: Culture, Economy and Conservation in Central Africa. Online edition, Oxford Academic, 28 Sept. 2023. 10.1093/oso/9780197746370.001.0001 (Accessed 28 Jan. 2025).

[10] Mahoney S. Eating Eden to Extinction: Understanding Africa’s Bushmeat Crisis. NRA hunters’ leadership forum [Internet], Sep 2022. Retrieved from: https://www.nrahlf.org/articles/2022/9/1/eating-eden-to-extinction-understanding-africa-s-bushmeat-crisis/ (Accessed 3/12/2024)

[11] Chaber A, Allebone-Webb S, Lignereux Y, Cunningham AA, & Rowcliffe JM. The scale of illegal meat importation from Africa to Europe via Paris. Conservation Letters, 2010; 3(5):317–321. 10.1111/j.1755-263X.2010.00121.x

[12] Nigeria Centre for Disease Control [Internet] Situation report. Lassa fever outbreak response, 2018. Available at: https://ncdc.gov.ng/themes/common/files/sitreps/57c8c2e9022f9cc34e470ba9b3078c91.pdf

[13] Fasina FO, Shittu A, Lazarus D, Tomori O, Simonsen L, Viboud C. et al. Transmission dynamics and control of Ebola virus disease outbreak in Nigeria, July to September 2014. Euro surveillance, 2014; 19(40): 20920. 10.2807/1560-7917.es2014.19.40.20920

[14] Grimes KEL, Ngoyi BF, Stolka KB, Hemingway-Foday JJ, Lubula L, Mossoko M, et al. Contextual, Social and Epidemiological Characteristics of the Ebola Virus Disease Outbreak in Likati Health Zone, Democratic Republic of the Congo, 2017. Frontiers in Public Health, 2020; 8:349. 10.3389/fpubh.2020.00349

[15] Dzotsi EK, Ohene SA, Asiedu-Bekoe F, Amankwa J, Sarkodie B, Adjabeng M et al. The first cases of Lassa fever in Ghana. Ghana Medical Journal, 2012; 46(3):166–70. PMID: 23661832; PMCID: PMC3645162.

[16] Haapala I & Probart C. Food safety knowledge, perceptions and behaviours among middle school students. J Nutr Educ Behaviour, 2004;36:71–76. doi: 10.1016/s1499-4046(06)60136-x

[17] [17] Lagos State Government [Internet]. Lagos Resilience Strategy, 2020. Available at: https://docslib.org/doc/3582298/lagos-resilience-strategy

[18] Braun V & Clarke V. Thematic analysis. In: Encyclopedia of Critical Psychology, pp. 1947–1952. Springer, 2014.

[19] Braun V & Clarke V. Using Thematic Analysis in Psychology. Qualitative Research in Psychology. 2006 Jul 21;3(2):77–101. 10.1191/1478088706qp063oa

[20] Cawthorn DM & Hoffman LC. The bushmeat and food security nexus: A global account of the contributions, conundrums and ethical collisions. Food Research International, 2015; 76(4):906–25. 10.1016/j.foodres.2015.03.025

[21] Yobo CM, Iponga DM, Coad L, & Ingram DJ. Contemporary wild meat hunting, consumption, and trade in Africa. African Journal of Ecology, 2022; 60(2):133–134. 10.1111/aje.13026

[22] Grace D, Bett B, Cook E, Lam S, MacMillan S, Masudi P., et al. Eating wild animals: Rewards, risks and recommendations. Nairobi, Kenya: ILRI, 2024. Available at: https://hdl.handle.net/10568/152246

[23] Robinson AL, Lee HJ, Kwon J, Todd E, Rodriguez FP, & Ryu D. Adequate Hand Washing and Glove Use Are Necessary to Reduce Cross-Contamination from Hands with High Bacterial Loads. Journal of food protection, 2016; 79(2):304–308. 10.4315/0362-028X.JFP-15-342

[24] Lues JFR & Tonder I. The occurrence of indicator bacteria on hands and aprons of food handlers in the delicatessen sections of a retail group. Food Control, 2007;18:326–332.

[25] Ali MM, Verrill L, & Zhang Y. Self-Reported Hand Washing Behaviors and Foodborne Illness: A Propensity Score Matching Approach. Journal of Food Protection, 2014; 77(3):352–8. 10.4315/0362-028X.JFP-13-286

[26] Adesola RO, Hossain D, Ogundijo OA, Idris I, Hamzat A, Bakre AA, et al. Challenges, health risks and recommendations on meat handling practices in Africa: a comprehensive review. Environmental Health Insights, 2024;18. doi:10.1177/11786302241301991

[27] Ogundipe OT, Atkinson PM, & Marsden SJ. Monitoring land use and land cover change in Lagos, Nigeria. International Journal of Applied Earth Observation and Geoinformation, 2016; 52:308–321.

[28] Akpan SN, van Hooft P, Happi AN, van Langevelde F, Buij R, Hassell JM, et al. Structure, Conservation, and Health Implications of Urban Wild Meat Value Chains: A Case Study of Lagos City, Nigeria. In press.

[29] Kurpiers LA, Schulte-Herbrüggen B, Ejotre I, & Reeder DM. Bushmeat and Emerging Infectious Diseases: Lessons from Africa. Problematic Wildlife, 2015 Sep 21:507–51. doi: 10.1007/978-3-319-22246-2_24. PMCID: PMC7123567.

[30] Bonwitt J, Dawson M, Kandeh M, Ansumana R., Sahr F, Brown H et al. Unintended consequences of the ‘bushmeat ban’ in West Africa during the 2013–2016 Ebola virus disease epidemic, Social Science & Medicine, 2018; 200:166–173. 10.1016/j.socscimed.2017.12.028

[31] Lâm S, Masudi SP, Nguyen HTT et al. Preventing mpox at its source: Using food safety and One Health strategies to address bushmeat practices. BMC Global Public Health, 2024; 2:69. 10.1186/s44263-024-00100-2

[32] Todd ECD, Michaels BS, Greig JD, Smith D, & Bartleson CA. Outbreaks where food workers have been implicated in the spread of foodborne disease. Part 8. Gloves as barriers to prevent contamination of food by workers. Journal of Food Protection, 2010; 73(9):1762–1773. 10.4315/0362-028X-73.9.1762

[33] Lobb, A. E. Consumer behavior and perceptions of food safety. In R. L. Scharff (Ed.), The Economics of Food Safety: The Case of Green Onions and Hepatitis A Outbreaks. Springer, 2019; 41–64.

[34] Mohan KR. Enhancing Food Safety: Understanding Consumer Awareness, Knowledge, and Concerns Regarding Food Contaminants. Indian Journal of Nutrition, 2024;11(1): 289.

[35] Odetokun IA, Borokinni BO, Bakare SB, Ghali-Mohammed I, Alhaji NB. A Cross-Sectional Survey of Consumers’ Risk Perception and Hygiene of Retail Meat: A Nigerian Study. Food Protection Trends, 2021; 41(3): 274–283.

[36] Woo PCY, Lau SKP, & Yuen K. Infectious diseases emerging from Chinese wet markets: zoonotic origins of severe respiratory viral infections. Current Opinion in Infectious Diseases, 2006; 19(5):401–407. 10.1097/01.qco.0000244043.08264.fc

[37] Olaoye OA, Onilude AA, & Idowu OA. Microbiological Profile of Goat Meat Inoculated with Lactic Acid Bacteria Cultures and Stored at 30°C for 7 days. Food Bioprocess Technology, 2011; 4:312–319. 10.1007/s11947-010-0343-3

[38] de Almeida A, Quaresma N, & Biosse E. The role of energy efficiency and renewable energies to accelerate sustainable energy access — a perspective case study of Mozambique. Energy Efficiency, 2022; 15(6): 36. 10.1007/s12053-022-10045-w

[39] Abernethy K & Obiang AM. Bushmeat in Gabon. Ministry of Water and Forests, Government of Gabon, 2010. Accessed at: http://hdl.handle.net/1893/26126

[40] Nielsen MR, Pouliot M, Meilby H, Smith-Hall C, & Angelsen A. Global patterns and determinants of the economic importance of wild meat. Biological Conservation, 2017; 215: 277–87. 10.1016/j.biocon.2017.08.036

[41] Mao Y, Ma P, Li T, Liu H, Zhao X, Liu S, et al. Flash heating process for efficient meat preservation. Nature Communications, 2024 May 8;15(1). 10.1038/s41467-024-47967-1

[42] Sahin S, Ulusoy HI, Alemdar S, Erdogan S, Agaoglu S. The Presence of Polycyclic Aromatic Hydrocarbons (PAHs) in Grilled Beef, Chicken and Fish by Considering Dietary Exposure and Risk Assessment. Food Sci Anim Resour, 2020; 40(5):675–688. doi: 10.5851/kosfa.2020.e43

[43] Lee JG, Kim SY, Moon JS, Kim SH, Kang DH, Yoon HJ. Effects of grilling procedures on levels of polycyclic aromatic hydrocarbons in grilled meats. Food Chemistry [Internet]. 2016 May 15; 199:632–8. https://pubmed.ncbi.nlm.nih.gov/26776018/

[44] Lu J, Zhang Y, Zhou H, Cai K, Xu B. A review of hazards in meat products: Multiple pathways, hazards and mitigation of polycyclic aromatic hydrocarbons. Food chemistry, 2024 Feb 1;138718–8. 10.1016/j.foodchem.2024.138718

[45] Lagos State Environmental Protection Agency [Internet]. Environmental-management-protection law, 2017. Available at: https://www.lasepa.gov.ng/documents/Environmental-Management-Protection-Law-2017-1.pdf

[46] Food and Agricultural Organization of the United Nations. Health Division: Meat & Meat Products. FAO FsAP, Rome, 2014.

[47] Bekker JL, Hoffman LC & Jooste PJ. Wildlife-associated zoonotic diseases in some southern African countries in relation to game meat safety: A review. Onderstepoort Journal of Veterinary Research, 2012; 79(1): 422. 10.4102/ojvr.v79i1.422

[48] Food and Agricultural Organization (FAO). Improvement of meat inspection system to scale up food safety and nutrition security in Ghana. FAO in Ghana, 2021.

[49] Tumelty L, Fa JE, Coad L, Friant S, Mbane J, Kamogne CT, et al. A systematic mapping review of links between handling wild meat and zoonotic diseases. One Health, 2023; 17:100637. 10.1016/j.onehlt.2023.100637

